# Validation of the PACER-PT: a measure of health-relevant perceptions of gentrification-related neighbourhood change

**DOI:** 10.64898/2026.11.03.26366862

**Authors:** Henrique Alves, Raquel Carvalheiro, Cláudia Jardim Santos, José Pedro Silva, Jana A. Hirsch, Yvonne L. Michael, Carla Moreira, Ana Isabel Ribeiro

## Abstract

**Objective:** Urban neighbourhoods across Europe are rapidly transforming through gentrification, with Portuguese cities particularly affected. Although the health impacts of these changes are increasingly recognised, validated instruments to assess perceptions of neighbourhood change remain scarce. This study adapted and validated a Portuguese version of the Perceptions About Change in Environment and Residents (PACER-PT) scale.

**Methods:** A cross-sectional online survey was conducted among 255 adults living for ≥5yrs in the Lisbon and Porto Metropolitan Areas. PACER was translated and culturally adapted to create PACER-PT. We evaluated psychometric properties using exploratory factor analysis (EFA), internal consistency, and convergent and criterion validity.

**Results:** EFA identified a three-factor structure explaining 47.5% of the variance: Built, social and economic changes, Positive attitude towards neighbourhood changes, and Displacement concerns and neighbourhood disruption. Internal consistency was acceptable (Cronbach’s alpha=0.70-0.78). Convergent/criterion validity were supported by significant associations with amount and pace of change, self-rated health, and quality of life. Younger and wealthier participants and those with greater residential satisfaction reported more favourable perceptions.

**Conclusion:** PACER-PT demonstrated it is a valid instrument for assessing gentrification-related neighbourhood changes in Portugal.

## INTRODUCTION

Decades of research have established neighbourhood environments as critical determinants of health outcomes [1]. Neighbourhoods are dynamic systems shaped by population movement, public and private investment, policy interventions, and broader socioeconomic processes [2]. Among these transformations, gentrification has emerged as one of the most widely studied forms of urban change. Gentrification involves an influx of residents with greater economic power than the existing population, accompanied by rising property values and the potential displacement of long-term, lower-income residents [3]. Consequently, gentrification can influence population health and health inequalities by transforming neighbourhoods [3].

Gentrification has traditionally been measured using objective census-based indicators, including changes in income, education, housing costs and demographic composition [4,5]. While such objective measures provide valuable contextual information, they fail to capture the lived experiences of residents undergoing neighbourhood transformation. Residents’ perceptions of neighbourhood change may represent an important mechanism linking urban change to health outcomes [6]. Such perceptions – residents’ subjective awareness and appraisal of changes occurring in their neighbourhood – may influence psychological stress, sense of belonging, social relationships, and housing security. By contrast, residents’ experiences encompass the broader ways in which neighbourhood transformation affects their everyday lives. Qualitative studies have provided important insights into these lived experiences [6–8], but their findings are typically limited in scalability and generalizability [9]. A key gap in the literature is therefore the lack of scalable measures of perceived neighbourhood change and of a clear conceptualization of how these perceptions may influence health [9].

Recognizing these limitations, Hirsch and colleagues developed the Perceptions About Change in Environment and Residents (PACER) scale to measure residents’ experiences of gentrification-related neighbourhood change relevant for health [9]. The development of PACER followed a rigorous multi-stage process including literature review, expert consultation, cognitive interviews, and pilot testing. The resulting instrument captured multiple dimensions of neighbourhood change, including built environment changes, social dynamics, affordability, and residents’ emotional responses to transformation. Psychometric evaluation demonstrated acceptable internal consistency and evidence of criterion-related validity across several domains.

PACER provides an important tool for measuring perceived gentrification-related neighbourhood change and has been developed, validated and applied in North American settings, including the original study in Philadelphia, its subsequent validation in MESA, and its application in the Canadian INTERACT study [9–11]. Its development incorporated input from the National Neighbourhood Indicators Partnership (NNIP), reflecting the challenges of capturing diverse experiences of neighbourhood change even within the United States [9].

Cross-national differences in housing systems, institutional arrangements, cultural contexts and historical patterns of urban development further reinforce the need for adaptation and validation. Although gentrification has a global reach, its drivers, manifestations and consequences vary across national and local contexts [6,12]. These differences highlight the importance of assessing whether PACER adequately captures residents’ experiences in settings beyond North America. In recent years, Portuguese cities, particularly Lisbon and Porto, have emerged as hotspots of gentrification, experiencing profound urban transformations shaped by rapid tourism growth, real estate investment, infrastructure development, and policy choices concerning rental regulation, foreign investment and tourism promotion [13,14]. These processes have raised growing public and academic concern regarding their social and health implications. Studies conducted in Porto Metropolitan Area [15–17] describe a complex scenario characterized by both negative and positive impacts, which disproportionately affect socially vulnerable groups. The most salient health-related consequences were negative, including adverse mental health outcomes associated with rising living costs, displacement, social disruption, exposure to environmental stressors, and reduced access to essential services. Nevertheless, some positive changes were also reported, including improvements in neighbourhood aesthetics, urban sanitation, economic opportunities, and opportunities for cultural exchange. Despite growing recognition of these processes and their societal and health impacts, quantitative research on perceived neighbourhood change in Portuguese contexts remains limited. Much of the existing research uses qualitative approaches, highlighting the need for standardised tools to assess residents’ perceptions of neighbourhood change consistently and at scale. The absence of validated instruments constrains large-scale epidemiological research on the health impacts of gentrification.

Under this background, the objective of this study was to adapt and validate the Portuguese version of the Perceptions About Change in Environment and Residents scale (PACER-PT-Perceções sobre Alterações na Área de Residência) for use in urban health research in Portugal. This included translation and cultural adaptation, followed by assessment of the psychometric properties and validity of PACER-PT.

## METHODS

### Sample and study design

This cross-sectional study was conducted in the Lisbon and Porto metropolitan areas, Portugal, between December 2025 and January 2026, within the HOT-Cities project (Health Outcomes of Touristification and Gentrification in Portuguese Cities: A Nationwide Survey), funded by Fundação “la Caixa” (FP25-1B113). The project investigates the health impacts of touristification and gentrification in Portuguese urban contexts to inform urban planning and public health strategies.

Data were collected through an online survey administered by Pureprofile using its established panel. Eligible participants were aged 18 years or older, fluent in Portuguese, and had lived in their current neighbourhood in either metropolitan area for at least five years. Neighbourhood was defined as the area surrounding participants’ homes, without a fixed geographical boundary, allowing respondents to use their own understanding of the term. Unlike the original PACER validation, which included participants with varying lengths of residence, this study focused on longer-term residents to ensure sufficient exposure to neighbourhood change.

The questionnaire included PACER-PT and questions characterising participants’ residential histories, housing situations, and sociodemographic characteristics. These covered duration of residence in the neighbourhood and current dwelling, housing tenure, intentions regarding future residence, reasons for choosing the neighbourhood, age, gender, educational attainment, employment status, household size, and household income. These variables were used to describe the sample and contextualise variation in scale responses.

Based on the recommended minimum ratio of 10 participants per item for scale validation [18], the 25-item scale required at least 250 valid responses. A total of 255 valid responses were obtained from a convenience sample. Comparison with the 2021 Census population of the Lisbon and Porto metropolitan areas (Supplementary File S1, Table 1) indicated a similar distribution of self-reported gender to the Census distribution of sex. However, younger and more highly educated individuals were overrepresented, while older adults and those with lower educational attainment were underrepresented.

**Table 1.**
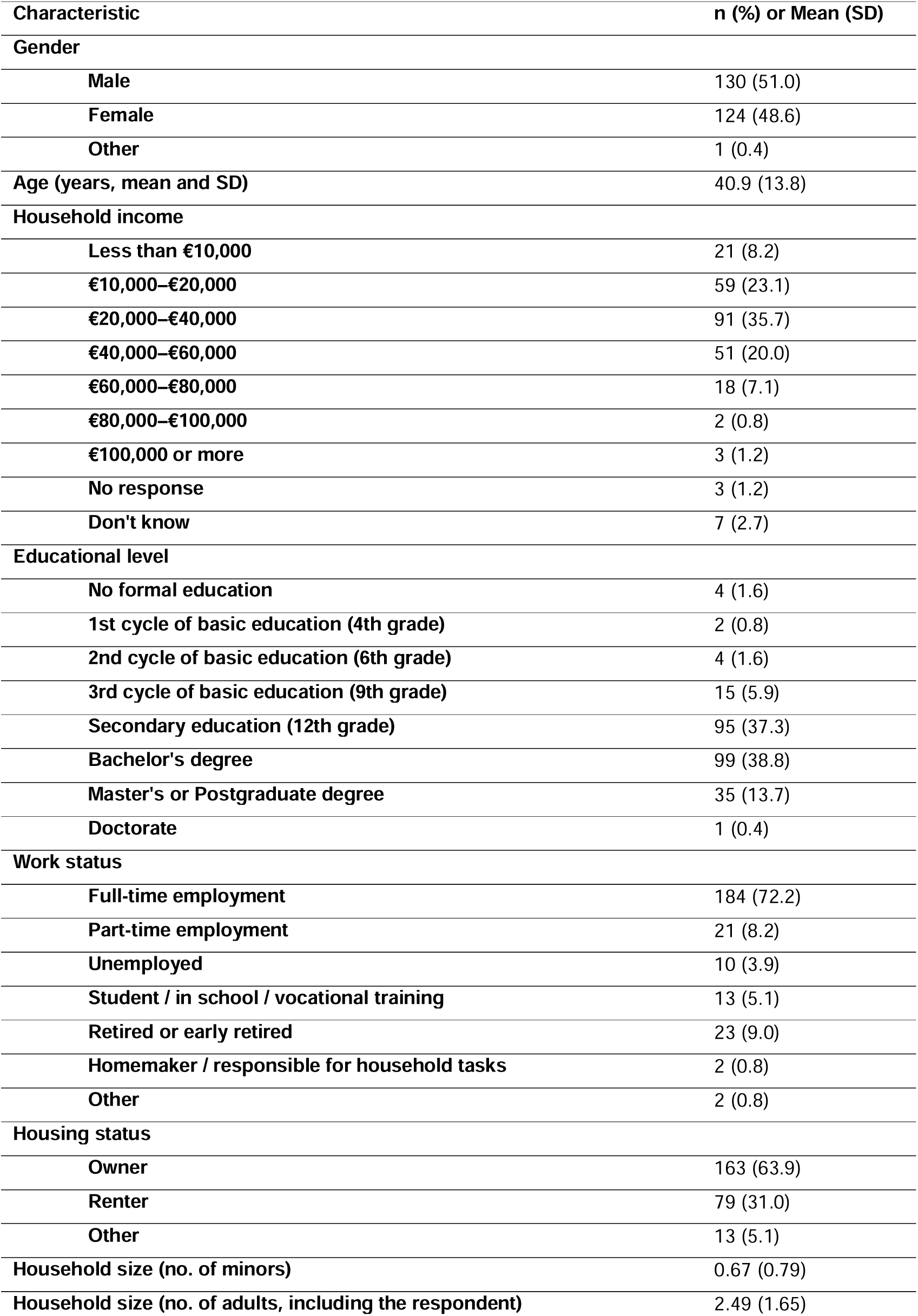
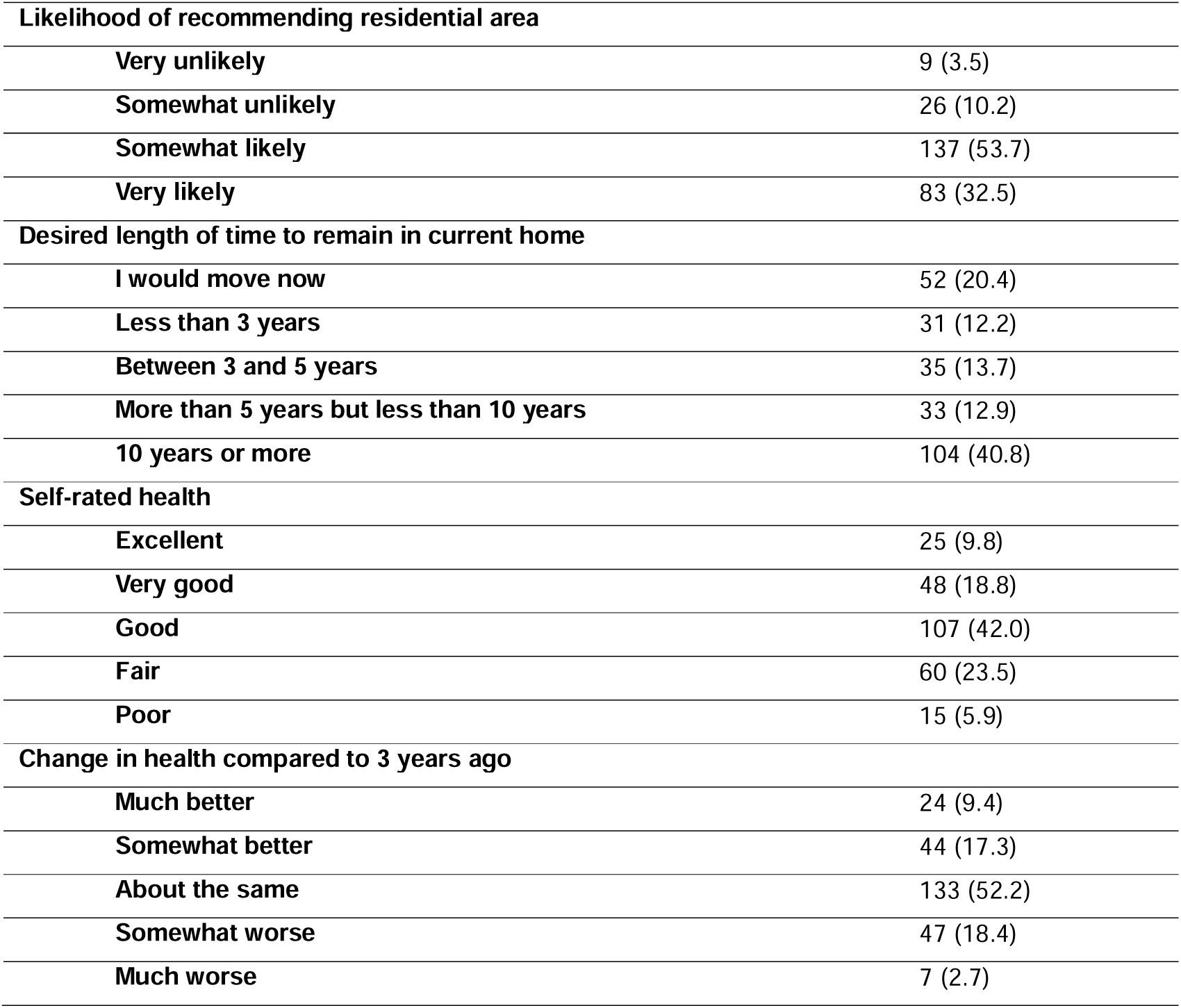
Sample characteristics (Porto and Lisbon Metropolitan Area, 2025-2026, n=255).

| Characteristic | **n (%) or Mean (SD)** |
|---|---|
| **Gender** |  |
| **Male** | 130 (51.0) |
| **Female** | 124 (48.6) |
| **Other** | 1 (0.4) |
| **Age (years, mean and SD)** | 40.9 (13.8) |
| **Household income** |  |
| **Less than €10,000** | 21 (8.2) |
| **€10,000–€20,000** | 59 (23.1) |
| **€20,000–€40,000** | 91 (35.7) |
| **€40,000–€60,000** | 51 (20.0) |
| **€60,000–€80,000** | 18 (7.1) |
| **€80,000–€100,000** | 2 (0.8) |
| **€100,000 or more** | 3 (1.2) |
| **No response** | 3 (1.2) |
| **Don't know** | 7 (2.7) |
| **Educational level** |  |
| **No formal education** | 4 (1.6) |
| **1st cycle of basic education (4th grade)** | 2 (0.8) |
| **2nd cycle of basic education (6th grade)** | 4 (1.6) |
| **3rd cycle of basic education (9th grade)** | 15 (5.9) |
| **Secondary education (12th grade)** | 95 (37.3) |
| **Bachelor's degree** | 99 (38.8) |
| **Master's or Postgraduate degree** | 35 (13.7) |
| **Doctorate** | 1 (0.4) |
| **Work status** |  |
| **Full-time employment** | 184 (72.2) |
| **Part-time employment** | 21 (8.2) |
| **Unemployed** | 10 (3.9) |
| **Student / in school / vocational training** | 13 (5.1) |
| **Retired or early retired** | 23 (9.0) |
| **Homemaker / responsible for household tasks** | 2 (0.8) |
| **Other** | 2 (0.8) |
| **Housing status** |  |
| **Owner** | 163 (63.9) |
| **Renter** | 79 (31.0) |
| **Other** | 13 (5.1) |
| **Household size (no. of minors)** | 0.67 (0.79) |
| **Household size (no. of adults, including the respondent)** | 2.49 (1.65) |
| **Likelihood of recommending residential area** |  |
| **Very unlikely** | 9 (3.5) |
| **Somewhat unlikely** | 26 (10.2) |
| **Somewhat likely** | 137 (53.7) |
| **Very likely** | 83 (32.5) |
| **Desired length of time to remain in current home** |  |
| **I would move now** | 52 (20.4) |
| **Less than 3 years** | 31 (12.2) |
| **Between 3 and 5 years** | 35 (13.7) |
| **More than 5 years but less than 10 years** | 33 (12.9) |
| **10 years or more** | 104 (40.8) |
| **Self-rated health** |  |
| **Excellent** | 25 (9.8) |
| **Very good** | 48 (18.8) |
| **Good** | 107 (42.0) |
| **Fair** | 60 (23.5) |
| **Poor** | 15 (5.9) |
| **Change in health compared to 3 years ago** |  |
| **Much better** | 24 (9.4) |
| **Somewhat better** | 44 (17.3) |
| **About the same** | 133 (52.2) |
| **Somewhat worse** | 47 (18.4) |
| **Much worse** | 7 (2.7) |

### Translation and cultural adaptation

The original PACER scale was translated and culturally adapted following established guidelines for instrument validation [19]. One bilingual native speaker/translator and a public health researcher fluent in English independently translated the original items, and a consensus version was produced. A back-translation into English was then performed by one bilingual native speaker unfamiliar with the original scale to ensure conceptual equivalence. To assess face validity, the research team reviewed the items for clarity, relevance, and cultural appropriateness. Particular attention was given to ensuring that terminology describing housing dynamics, neighbourhood change, commercial activities, and social relationships reflected Portuguese linguistic conventions and urban realities. The pre-final version was pilot-tested with a small sample from the target population to verify comprehensibility (n=10).

### Measures

As in the original PACER scale, the Portuguese version (PACER-PT) was used to assess residents’ perceptions of neighbourhood change occurring over the previous three to five years. The PACER-PT scale is provided in the supplementary material (Supplementary File S2).

The original PACER questionnaire comprises nine items assessing the perceived occurrence of neighbourhood changes. Responses to these items were coded as follows: “It does not happen” (−1), “Unsure” (0), “It happens a little” (1), and “It happens a lot” (2). Thus, “Unsure” was assigned an intermediate numerical value between “It does not happen” and “It happens a little”. A further ten items assess emotional and cognitive responses to neighbourhood change using a five-point agreement scale. In the final survey analysis, responses were coded from −2 (“Strongly disagree”) to +2 (“Strongly agree”), with 0 corresponding to “Neither agree nor disagree”. “Not applicable” responses were also coded as 0. As done by Hirsch and colleagues, four of the items – signalled by (R) in Table 2 - were reverse-coded [9]. The items cover changes commonly associated with gentrification, including housing and commercial transformation, population change, neighbourhood tensions, belonging, trust and displacement concerns [9].

**Table 2.**
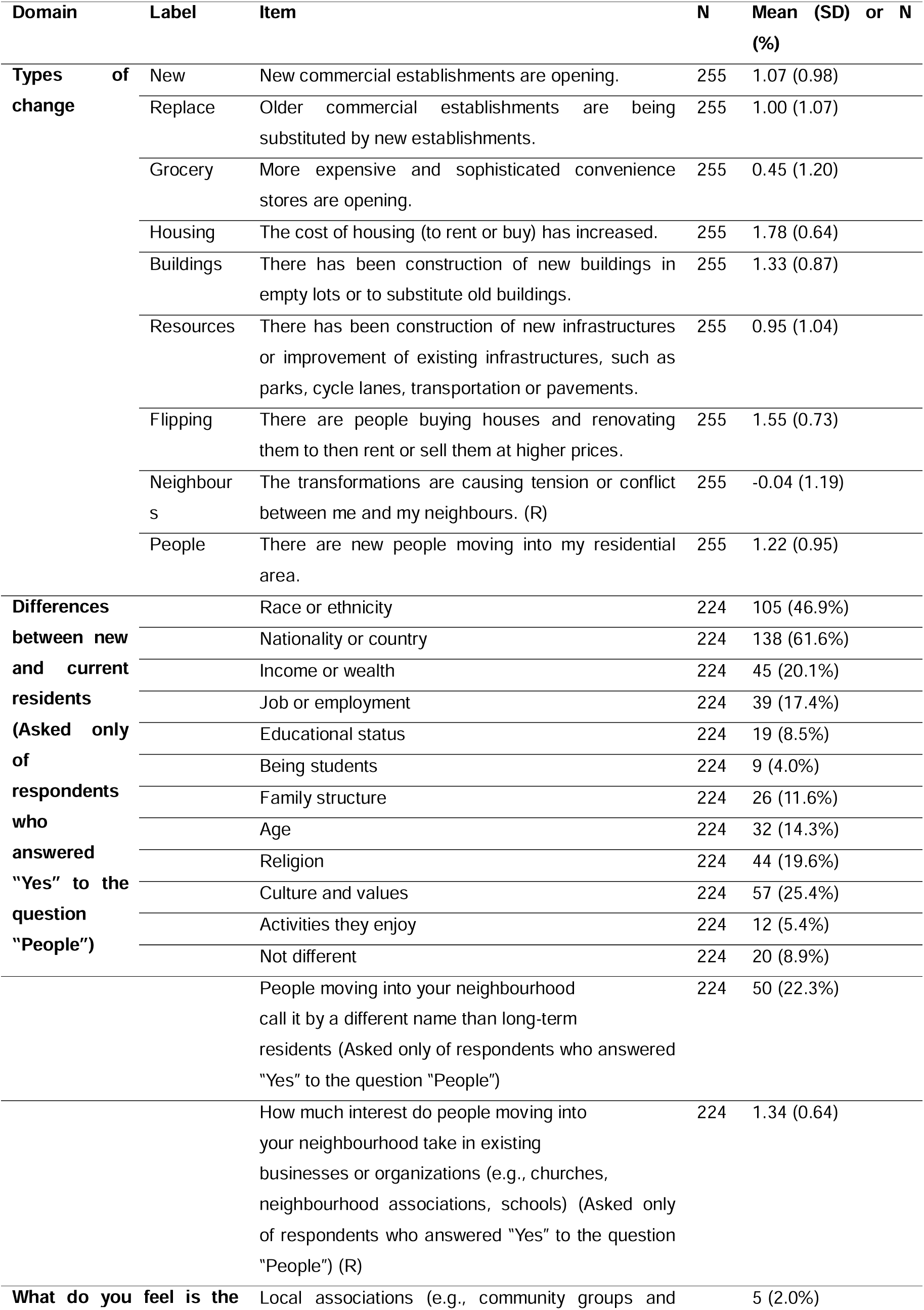
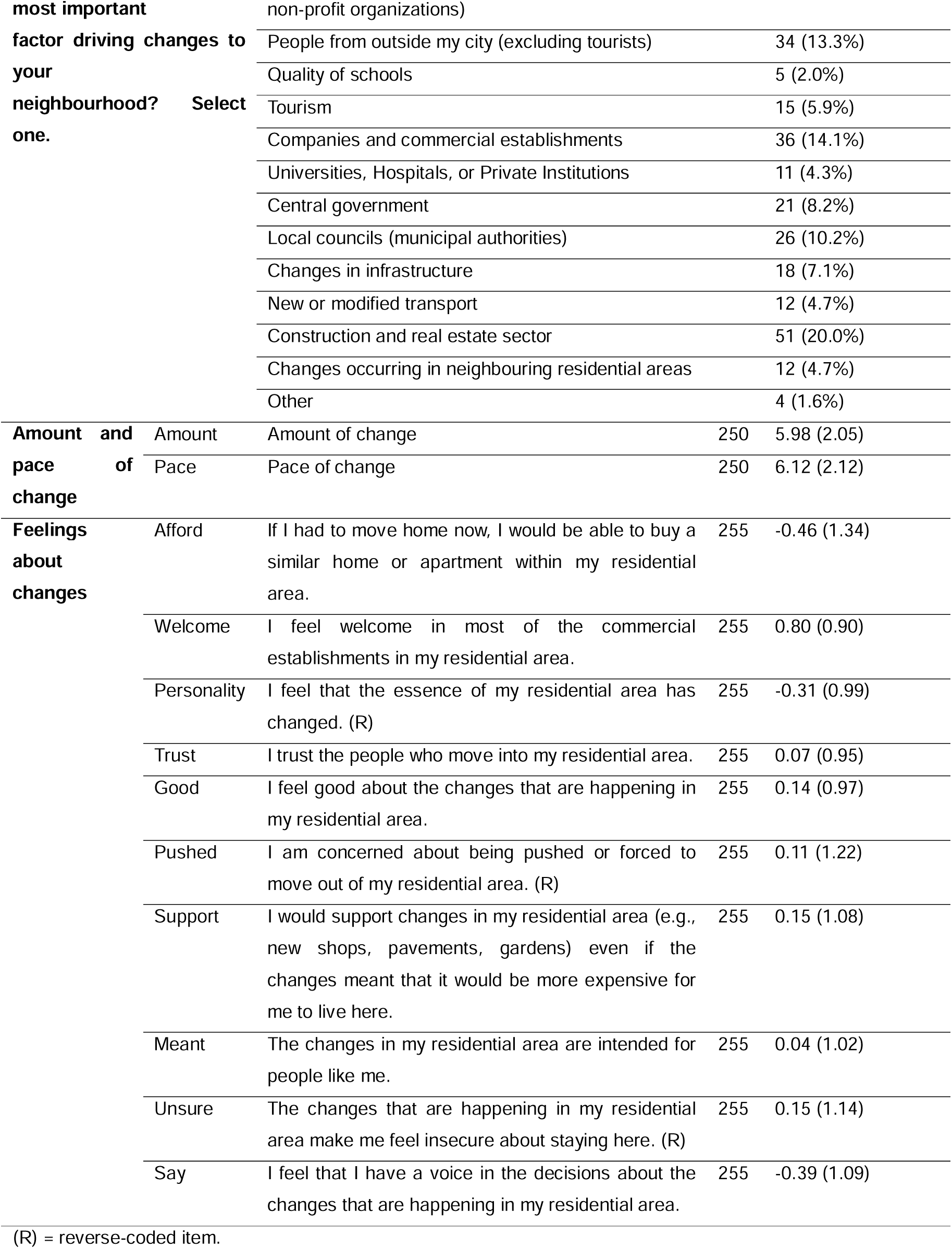
PACER-PT items and descriptive statistics (Porto and Lisbon Metropolitan Area, 2025-2026, n=255).

| Domain | Label | Item | N | Mean (SD) or N (%) |
|---|---|---|---|---|
| **Types of change** | New | New commercial establishments are opening. | 255 | 1.07 (0.98) |
|  | Replace | Older commercial establishments are being substituted by new establishments. | 255 | 1.00 (1.07) |
|  | Grocery | More expensive and sophisticated convenience stores are opening. | 255 | 0.45 (1.20) |
|  | Housing | The cost of housing (to rent or buy) has increased. | 255 | 1.78 (0.64) |
|  | Buildings | There has been construction of new buildings in empty lots or to substitute old buildings. | 255 | 1.33 (0.87) |
|  | Resources | There has been construction of new infrastructures or improvement of existing infrastructures, such as parks, cycle lanes, transportation or pavements. | 255 | 0.95 (1.04) |
|  | Flipping | There are people buying houses and renovating them to then rent or sell them at higher prices. | 255 | 1.55 (0.73) |
|  | Neighbours | The transformations are causing tension or conflict between me and my neighbours. (R) | 255 | -0.04 (1.19) |
|  | People | There are new people moving into my residential area. | 255 | 1.22 (0.95) |
| **Differences between new and current residents (Asked only of respondents who answered "Yes" to the question "People")** |  | Race or ethnicity | 224 | 105 (46.9%) |
|  |  | Nationality or country | 224 | 138 (61.6%) |
|  |  | Income or wealth | 224 | 45 (20.1%) |
|  |  | Job or employment | 224 | 39 (17.4%) |
|  |  | Educational status | 224 | 19 (8.5%) |
|  |  | Being students | 224 | 9 (4.0%) |
|  |  | Family structure | 224 | 26 (11.6%) |
|  |  | Age | 224 | 32 (14.3%) |
|  |  | Religion | 224 | 44 (19.6%) |
|  |  | Culture and values | 224 | 57 (25.4%) |
|  |  | Activities they enjoy | 224 | 12 (5.4%) |
|  |  | Not different | 224 | 20 (8.9%) |
|  |  | People moving into your neighbourhood<br>call it by a different name than long-term<br>residents (Asked only of respondents who answered<br>"Yes" to the question "People") | 224 | 50 (22.3%) |
|  |  | How much interest do people moving into<br>your neighbourhood take in existing<br>businesses or organizations (e.g., churches,<br>neighbourhood associations, schools) (Asked only of respondents who answered "Yes" to the question "People") (R) | 224 | 1.34 (0.64) |
| **What do you feel is the most important factor driving changes to your neighbourhood? Select one.** |  | Local associations (e.g., community groups and non-profit organizations) |  | 5 (2.0%) |
|  |  | People from outside my city (excluding tourists) |  | 34 (13.3%) |
|  |  | Quality of schools |  | 5 (2.0%) |
|  |  | Tourism |  | 15 (5.9%) |
|  |  | Companies and commercial establishments |  | 36 (14.1%) |
|  |  | Universities, Hospitals, or Private Institutions |  | 11 (4.3%) |
|  |  | Central government |  | 21 (8.2%) |
|  |  | Local councils (municipal authorities) |  | 26 (10.2%) |
|  |  | Changes in infrastructure |  | 18 (7.1%) |
|  |  | New or modified transport |  | 12 (4.7%) |
|  |  | Construction and real estate sector |  | 51 (20.0%) |
|  |  | Changes occurring in neighbouring residential areas |  | 12 (4.7%) |
|  |  | Other |  | 4 (1.6%) |
| **Amount and pace of change** | Amount | Amount of change | 250 | 5.98 (2.05) |
|  | Pace | Pace of change | 250 | 6.12 (2.12) |
| **Feelings about changes** | Afford | If I had to move home now, I would be able to buy a similar home or apartment within my residential area. | 255 | -0.46 (1.34) |
|  | Welcome | I feel welcome in most of the commercial establishments in my residential area. | 255 | 0.80 (0.90) |
|  | Personality | I feel that the essence of my residential area has changed. (R) | 255 | -0.31 (0.99) |
|  | Trust | I trust the people who move into my residential area. | 255 | 0.07 (0.95) |
|  | Good | I feel good about the changes that are happening in my residential area. | 255 | 0.14 (0.97) |
|  | Pushed | I am concerned about being pushed or forced to move out of my residential area. (R) | 255 | 0.11 (1.22) |
|  | Support | I would support changes in my residential area (e.g., new shops, pavements, gardens) even if the changes meant that it would be more expensive for me to live here. | 255 | 0.15 (1.08) |
|  | Meant | The changes in my residential area are intended for people like me. | 255 | 0.04 (1.02) |
|  | Unsure | The changes that are happening in my residential area make me feel insecure about staying here. (R) | 255 | 0.15 (1.14) |
|  | Say | I feel that I have a voice in the decisions about the changes that are happening in my residential area. | 255 | -0.39 (1.09) |

(R) = reverse-coded item.

The Portuguese translation and cultural adaptations made to these items are described below. During cultural adaptation, particular attention was given to terminology relating to housing tenure, commercial change and neighbourhood governance. Wording and examples were modified where necessary to ensure their relevance and comprehensibility in the Portuguese context, while preserving the intended meaning of the original items. In addition to these items, contextual indicators were included to measure the perceived amount and pace of neighbourhood change (questions 13, 14). Participants rated the overall magnitude of neighbourhood transformation and the speed at which changes had occurred using numeric scales ranging from 1 to 10. These indicators were used to support the evaluation of convergent validity.

Questions 10, 11, 12, and 15 were included as additional items to capture complementary dimensions of perceived neighbourhood change not covered by the core PACER items. Specifically, these questions assessed perceived demographic and socioeconomic differences between long-term residents and newcomers, perceived changes in neighbourhood naming practices, newcomers’ engagement with existing local establishments and organisations, and residents’ perceptions of the primary drivers of neighbourhood transformation, including tourism.

Moreover, health-related measures were included to assess general health and quality of life. Participants rated their overall quality of life on a five-point scale ranging from “Very poor” to “Very good”. Self-rated general health was assessed using a single item with five response options: “Excellent”, “Very good”, “Good”, “Fair”, and “Poor”. Perceived change in health status was also evaluated by asking participants to compare their current general health with that of three years earlier, with response options ranging from much better to much worse. These measures were used to examine criterion validity between PACER-PT scores and health-related outcomes.

### Statistical analysis

All statistical analyses were conducted following established procedures for psychometric validation of measurement instruments [19] and followed the steps of the authors of the original PACER scale [9].

Descriptive statistics were calculated for all PACER-PT items, including means and standard deviations, to examine response distributions and identify potential floor and ceiling effects.

Exploratory Factor Analysis (EFA) was conducted to examine the dimensional structure of the PACER-PT scale and identify latent constructs underlying patterns of responses. The EFA included the 19 core PACER-PT items (nine types of change and ten feelings about change); the complementary items were analysed descriptively. The suitability of the data for factor analysis was assessed using the Kaiser-Meyer-Olkin (KMO) measure of sampling adequacy and Bartlett’s test of sphericity. Factor extraction was performed using principal axis factoring. The number of factors to retain was determined based on eigenvalues greater than 1 (Kaiser criterion [20]), inspection of the scree plot, and the interpretability of the resulting factor structure. An oblique rotation (*oblimin* with kaiser normalization) was applied, as correlations between factors were expected. Items with factor loadings ≥0.40 were considered significant contributors to a factor. Items exhibiting cross-loadings ≥0.30 on more than one factor were examined carefully, and decisions regarding item retention were based on both statistical and theoretical considerations. Several alternative models were examined, and the final solution was selected based on statistical performance and theoretical plausibility.

Internal consistency acceptability was evaluated using Cronbach’s alpha coefficients calculated separately for each identified factor. Values of α ≥ 0.70 were considered indicative of acceptable internal consistency [21]. Subscale scores were calculated as unweighted averages of items loading onto each factor.

Convergent validity was assessed by calculating Spearman correlation coefficients between PACER-PT subscale scores and contextual indicators of perceived neighbourhood change, specifically perceived amount and pace of change (“On a scale of 1-10, with 10 being the most and 1 being the least, overall how much change has happened in your neighbourhood during the last three to five years?” and “On a scale of 1-10, with 10 being the fastest and 1 being the slowest, how quickly have changes been happening in your neighbourhood during the last three to five years?”).

Additional Spearman correlation coefficients were calculated between PACER-PT subscale scores and selected indicators of perceived quality of life and self-rated health to further assess criterion validity. All statistical analyses were conducted using SPSS 30.0.0.0, and statistical significance was set at p < 0.05.

Mean PACER-PT factor scores were compared across participants’ sociodemographic and contextual characteristics using independent-samples T test or one-way analysis of variance (ANOVA). To ensure adequate group sizes, several sociodemographic and contextual variables were aggregated into broader categories.

## RESULTS

### Sample characteristics

Sample characteristics are shown in Table 1. The sample included 255 participants (51.0% male), with a mean age of 40.9 years (SD=13.8). Most participants reported household incomes between €20,000 and €40,000 (35.7%), had completed secondary (37.3%) or bachelor’s level education (38.8%), were employed full-time (72.2%), and owned their homes (63.9%). Over half were somewhat likely to recommend their residential area (53.7%), and 40.8% intended to remain in their current home for at least 10 years. Most participants rated their health as good or very good (60.8%), with 52.2% reporting no change in health compared with three years earlier.

Regarding the PACER-PT questions, descriptive analyses, depicted in Table 2, indicated that respondents reported varying degrees of perceived neighbourhood transformation across multiple domains. Regarding the types of change, the highest mean scores were observed for items related to housing cost increases (Housing, mean = 1.78, SD = 0.64) and housing market activity (Flipping, mean = 1.55, SD = 0.73), while the lowest mean score was reported for items reflecting the opening of expensive and sophisticated stores (Grocery, mean = 0.45, SD = 1.20) and tension and conflict between neighbours (Neighbours, mean = - 0.04, SD = 1.19).

With respect to feelings about changes, the highest mean score was observed for feeling welcome in commercial establishments (Welcome, mean = 0.80, SD = 0.90), whereas the lowest mean score was found for perceived ability to afford housing within the neighbourhood (Afford, mean = −0.46, SD = 1.34). Intermediate mean scores (around 0 on average) were observed for items related to support for neighbourhood change (Support, mean = 0.15, SD = 1.08), trust in newcomers (Trust, mean = 0.07, SD = 0.95), and displacement concerns (Pushed, mean = 0.11, SD = 1.22).

### Exploratory factor analysis

Exploratory Factor Analysis (EFA) was conducted to examine the dimensional structure of the PACER-PT scale. Kaiser-Meyer-Olkin (KMO) measure of sampling adequacy was 0.774, indicating good adequacy for factor analysis. Bartlett’s test of sphericity was significant (χ²(171)

= 1399.025, p < 0.001), supporting the suitability of the data for factor analysis.

The final factor solution supported a three-factor structure, which explained approximately 47.5% of the total variance in the data. The eigenvalues associated with the three retained factors were 3.70, 3.18, and 2.15, respectively. The cumulative variance explained by the three factors reached 19.5%, 36.2%, and 47.5% at each successive step, indicating that the selected structure captured a substantial proportion of the variance in responses.

Table 3 shows the factor loadings of the final solution. The first factor captured observable changes in the built, social and economic environment. These items included the construction of new buildings, property renovation activities, increases in housing costs, the arrival of new residents, and commercial transformation. This factor was interpreted as representing “Built, social and economic changes”, reflecting structural shifts associated with gentrification processes.

**Table 3.** Internal consistency, dimensionality, and criterion-related validity of PACER-PT using exploratory factor analysis with principal axis factoring (Porto and Lisbon Metropolitan Area, 2025-2026, n=255).

|  | **Factor 1** | **Factor 2** | **Factor 3** |
|---|---|---|---|
|  | **Built, social and economic changes** | **Positive attitude towards neighbourhood changes** | **Displacement concerns and neighbourhood disruption** |
| **Question and factor loading** | Flipping (0.687)<br>Housing (0.647)<br>Buildings (0.628)<br>People (0.576)<br>New (0.544)<br>Resources (0.453)<br>Grocery (0.430)<br>Replace (0.400) | Say (0.706)<br>Good (0.682)<br>Meant (0.641)<br>Trust (0.640)<br>Afford (0.527)<br>Support (0.505)<br>Welcome (0.323) | Unsure (0.726)<br>Pushed (0.68)<br>Personality (0.526)<br>Neighbours (0.443) |
| **Eigenvalue** | 3.70 | 3.18 | 2.15 |
| **Cum. Var. explained (%)** | 19.5 | 36.2 | 47.5 |
| **Cronbach's alpha** | 0.77 | 0.78 | 0.70 |
| **Correl with factor 1 (rho)** | ---- | 0.084 | **- 0.268**<sup>\*\*\*</sup> |
| **Correl with factor 2 (rho)** | --- | --- | 0.108 |
| **Correl. with Amount (rho)** | **0.317**<sup>\*\*\*</sup> | **- 0.145**<sup>\*</sup> | **- 0.334**<sup>\*\*\*</sup> |
| **Correl. with Pace (rho)** | **0.337**<sup>\*\*\*</sup> | - 0.085 | **- 0.257**<sup>\*\*\*</sup> |
| **Corr. with Quality of Life (rho)** | **0.153**<sup>\*</sup> | **0.272**<sup>\*\*\*</sup> | **0.139**<sup>\*</sup> |
| **Corr. with Self-rated Health (rho)** | 0.083 | **0.192**<sup>\*\*</sup> | 0.034 |
| **Corr. with Self-rated Health Change (rho)** | 0.095 | **0.314**<sup>\*\*\*</sup> | - 0.011 |

The second factor included items reflecting residents’ positive attitudes toward neighbourhood change. This dimension included items related to feeling welcome in new businesses, trusting new residents, feeling that neighbourhood changes were meant for people like oneself, supporting neighbourhood improvements, and perceiving that one had a voice in neighbourhood transformation processes. This factor was interpreted as representing positive responses to neighbourhood change and was therefore labelled “Positive attitude towards neighbourhood changes”.

The third factor included items reflecting uncertainty and perceived risk related to neighbourhood transformation. Items loading onto this factor included concerns about displacement, fear of being pushed out of the neighbourhood, perceptions of changes in neighbourhood character, and reported tensions between neighbours. This factor was interpreted as representing “Displacement concerns and neighbourhood disruption”, although due to the reverse-coding of the corresponding items, the greater its value, the smaller these fears and concerns are perceived.

### Reliability analysis

Internal consistency was acceptable across all factors, with Cronbach’s alpha values of 0.77 (Built, social and economic changes), 0.78 (Positive attitude towards neighbourhood changes), and 0.70 (Displacement concerns and neighbourhood disruption).

### Convergent and criterion-related validity

Correlation analyses provided further support for convergent and criterion validity (Table 3). The Built, social and economic changes factor showed significant positive correlations with perceived amount of change (ρ=0.317, p<0.001) and pace of change (ρ=0.337, p<0.001), indicating that higher perceptions of structural and economic transformation were associated with greater perceived intensity of neighbourhood change.

Consistent with these findings, the “Displacement concerns and neighbourhood disruption” factor was negatively associated with the perceived amount (ρ = − 0.334, p < 0.001) and pace of neighbourhood change (ρ = − 0.257, p < 0.001). As higher factor scores indicate fewer concerns, these associations suggest that greater perceived change and a faster perceived pace of change were associated with greater concerns about displacement and neighbourhood uncertainty. The “Positive attitude towards neighbourhood changes” factor showed a weak but significant association with perceived amount of change (ρ = −0.145, p<0.05) suggesting that positive perceptions about the changes decline with perceived intensity of neighbourhood change.

The “Positive attitude towards neighbourhood changes” factor was positively correlated with quality of life (ρ = 0.272, p < 0.001), self-rated health (ρ = 0.192, p < 0.01), and perceived changes in health status (ρ = 0.314, p < 0.001).The “Displacement concerns and neighbourhood disruption” and “Built, social and economic changes” factors were also positively, but less strongly, associated with quality of life (ρ=0.139, p<0.05 and ρ=0.153, p<0.05, respectively), although the associations with self-rated health and perceived changes in health status did not reach significance, suggesting that they had a more limited effect in perceived health outcomes.

### PACER-PT factor scores according socioeconomic and demographic factors

Table 4 shows the mean PACER-PT factor scores according to sociodemographic and contextual characteristics. Overall, Factor 2 (Positive attitude towards neighbourhood changes) showed the greatest variation across groups. Significant differences were observed according to age (*p* = 0.016), household income (*p* = 0.033), likelihood of recommending the residential area (*p* < 0.001), and desired length of time to remain in the current home (*p* < 0.001). Younger participants (<45 years) and those with higher household income (≥ €60.000) tended to report more positive attitude towards neighbourhood changes than the others. Participants who were very likely to recommend their residential area and those intending to remain in their home for longer periods also reported significantly more positive attitudes towards neighbourhood changes.

**Table 4.**
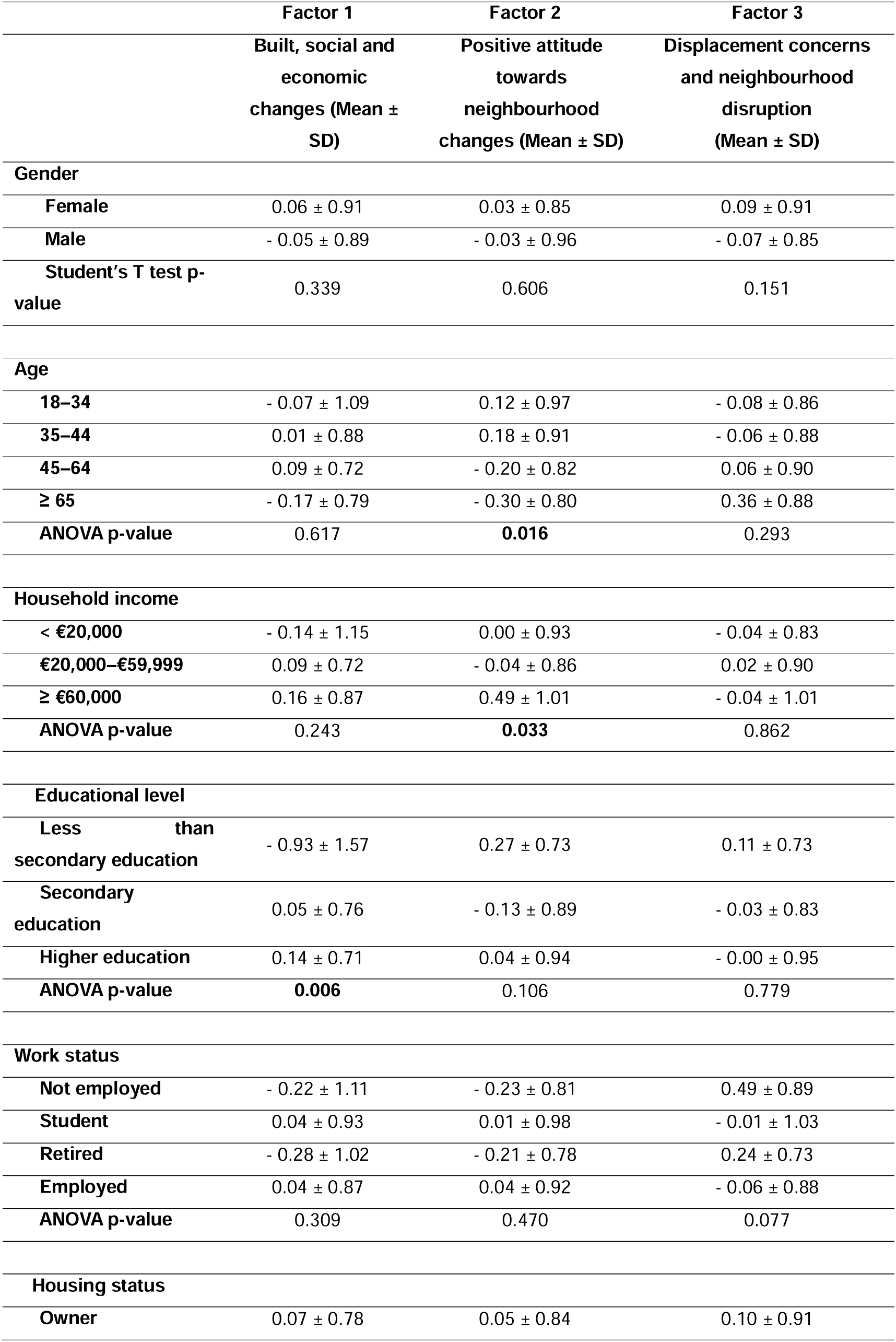
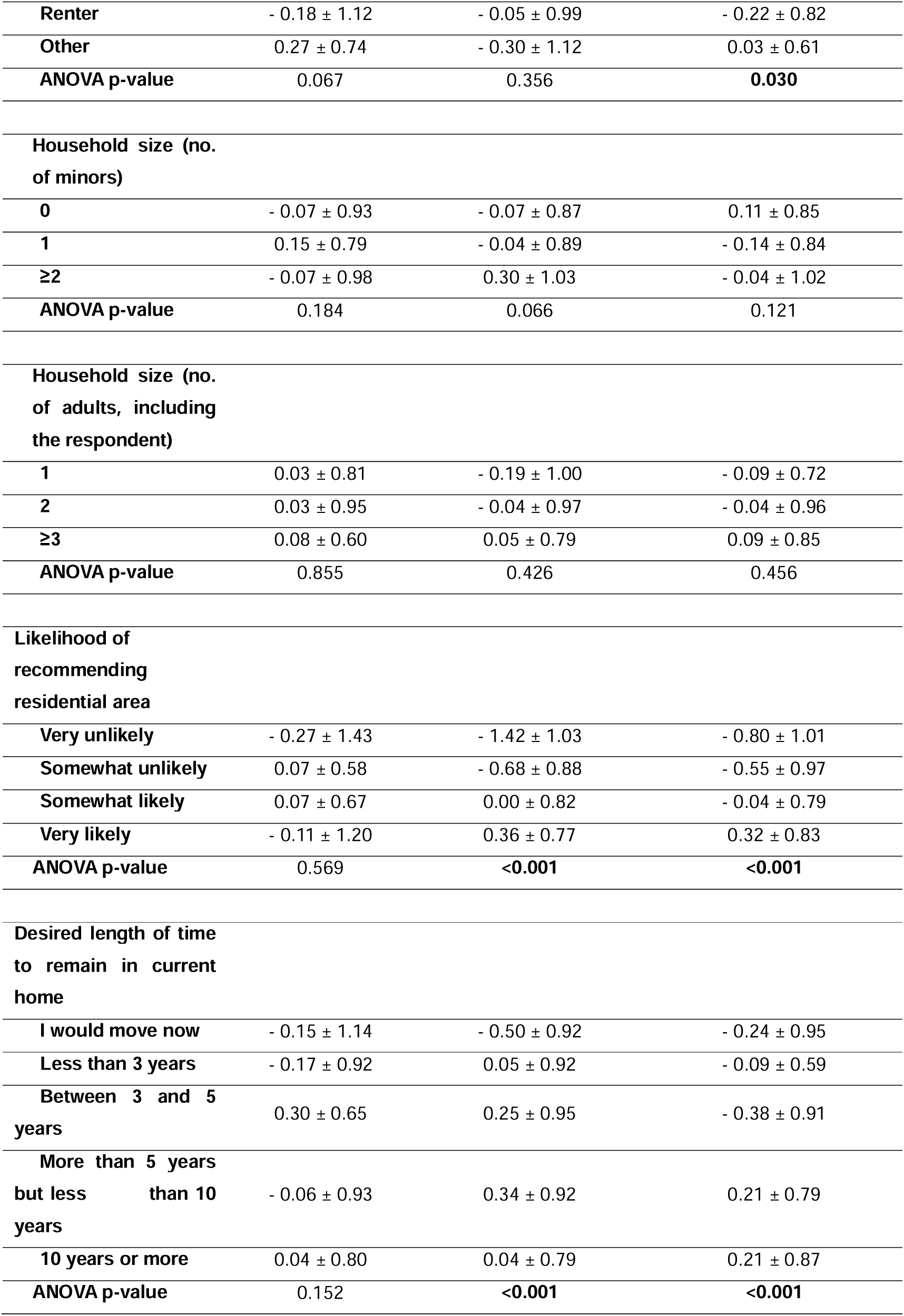
Mean PACER-PT factor scores according to sociodemographic and contextual characteristics (Porto and Lisbon Metropolitan Area, 2025-2026, n=255).

|  | **Factor 1** | **Factor 2** | **Factor 3** |
|---|---|---|---|
|  | **Built, social and economic changes (Mean ± SD)** | **Positive attitude towards neighbourhood changes (Mean ± SD)** | **Displacement concerns and neighbourhood disruption (Mean ± SD)** |
| **Gender** |  |  |  |
| **Female** | 0.06 ± 0.91 | 0.03 ± 0.85 | 0.09 ± 0.91 |
| **Male** | - 0.05 ± 0.89 | - 0.03 ± 0.96 | - 0.07 ± 0.85 |
| **Student's T test p-value** | 0.339 | 0.606 | 0.151 |
| **Age** |  |  |  |
| **18–34** | - 0.07 ± 1.09 | 0.12 ± 0.97 | - 0.08 ± 0.86 |
| **35–44** | 0.01 ± 0.88 | 0.18 ± 0.91 | - 0.06 ± 0.88 |
| **45–64** | 0.09 ± 0.72 | - 0.20 ± 0.82 | 0.06 ± 0.90 |
| **≥ 65** | - 0.17 ± 0.79 | - 0.30 ± 0.80 | 0.36 ± 0.88 |
| **ANOVA p-value** | 0.617 | **0.016** | 0.293 |
| **Household income** |  |  |  |
| **< €20,000** | - 0.14 ± 1.15 | 0.00 ± 0.93 | - 0.04 ± 0.83 |
| **€20,000–€59,999** | 0.09 ± 0.72 | - 0.04 ± 0.86 | 0.02 ± 0.90 |
| **≥ €60,000** | 0.16 ± 0.87 | 0.49 ± 1.01 | - 0.04 ± 1.01 |
| **ANOVA p-value** | 0.243 | **0.033** | 0.862 |
| **Educational level** |  |  |  |
| **Less than secondary education** | - 0.93 ± 1.57 | 0.27 ± 0.73 | 0.11 ± 0.73 |
| **Secondary education** | 0.05 ± 0.76 | - 0.13 ± 0.89 | - 0.03 ± 0.83 |
| **Higher education** | 0.14 ± 0.71 | 0.04 ± 0.94 | - 0.00 ± 0.95 |
| **ANOVA p-value** | **0.006** | 0.106 | 0.779 |
| **Work status** |  |  |  |
| **Not employed** | - 0.22 ± 1.11 | - 0.23 ± 0.81 | 0.49 ± 0.89 |
| **Student** | 0.04 ± 0.93 | 0.01 ± 0.98 | - 0.01 ± 1.03 |
| **Retired** | - 0.28 ± 1.02 | - 0.21 ± 0.78 | 0.24 ± 0.73 |
| **Employed** | 0.04 ± 0.87 | 0.04 ± 0.92 | - 0.06 ± 0.88 |
| **ANOVA p-value** | 0.309 | 0.470 | 0.077 |
| **Housing status** |  |  |  |
| **Owner** | 0.07 ± 0.78 | 0.05 ± 0.84 | 0.10 ± 0.91 |
| **Renter** | - 0.18 ± 1.12 | - 0.05 ± 0.99 | - 0.22 ± 0.82 |
| **Other** | 0.27 ± 0.74 | - 0.30 ± 1.12 | 0.03 ± 0.61 |
| **ANOVA p-value** | 0.067 | 0.356 | **0.030** |
| **Household size (no. of minors)** |  |  |  |
| **0** | - 0.07 ± 0.93 | - 0.07 ± 0.87 | 0.11 ± 0.85 |
| **1** | 0.15 ± 0.79 | - 0.04 ± 0.89 | - 0.14 ± 0.84 |
| **≥2** | - 0.07 ± 0.98 | 0.30 ± 1.03 | - 0.04 ± 1.02 |
| **ANOVA p-value** | 0.184 | 0.066 | 0.121 |
| **Household size (no. of adults, including the respondent)** |  |  |  |
| **1** | 0.03 ± 0.81 | - 0.19 ± 1.00 | - 0.09 ± 0.72 |
| **2** | 0.03 ± 0.95 | - 0.04 ± 0.97 | - 0.04 ± 0.96 |
| **≥3** | 0.08 ± 0.60 | 0.05 ± 0.79 | 0.09 ± 0.85 |
| **ANOVA p-value** | 0.855 | 0.426 | 0.456 |
| **Likelihood of recommending residential area** |  |  |  |
| **Very unlikely** | - 0.27 ± 1.43 | - 1.42 ± 1.03 | - 0.80 ± 1.01 |
| **Somewhat unlikely** | 0.07 ± 0.58 | - 0.68 ± 0.88 | - 0.55 ± 0.97 |
| **Somewhat likely** | 0.07 ± 0.67 | 0.00 ± 0.82 | - 0.04 ± 0.79 |
| **Very likely** | - 0.11 ± 1.20 | 0.36 ± 0.77 | 0.32 ± 0.83 |
| **ANOVA p-value** | 0.569 | **<0.001** | **<0.001** |
| **Desired length of time to remain in current home** |  |  |  |
| **I would move now** | - 0.15 ± 1.14 | - 0.50 ± 0.92 | - 0.24 ± 0.95 |
| **Less than 3 years** | - 0.17 ± 0.92 | 0.05 ± 0.92 | - 0.09 ± 0.59 |
| **Between 3 and 5 years** | 0.30 ± 0.65 | 0.25 ± 0.95 | - 0.38 ± 0.91 |
| **More than 5 years but less than 10 years** | - 0.06 ± 0.93 | 0.34 ± 0.92 | 0.21 ± 0.79 |
| **10 years or more** | 0.04 ± 0.80 | 0.04 ± 0.79 | 0.21 ± 0.87 |
| **ANOVA p-value** | 0.152 | **<0.001** | **<0.001** |

The only significant association found for Factor 1 (Built, social and economic changes) was relative to educational level (*p* = 0.006), where participants who did not complete secondary education reported fewer changes in their neighbourhoods than participants with secondary or higher education.

For Factor 3 (Displacement concerns and neighbourhood disruption), significant differences were observed for housing status (*p* = 0.030), likelihood of recommending the residential area (*p* < 0.001) and desired length of time to remain in the current home (*p* < 0.001). Homeowners reported fewer concerns about displacement and neighbourhood disruption than renters. The same happened with those who were very likely to recommend their neighbourhood and those desiring to remain in their current residence for longer periods.

## DISCUSSION

This study successfully translated and culturally adapted the PACER questionnaire into Portuguese and demonstrated that the resulting Portuguese version (PACER-PT) has satisfactory psychometric properties. Overall, the findings support the validity and reliability of the adapted instrument for assessing perceptions of gentrification-related neighbourhood change in Portugal. The PACER-PT demonstrated a coherent three-factor structure with acceptable internal consistency, meaningful associations with neighbourhood change and health-related outcomes, and the ability to discriminate between groups expected to differ in their perceptions of neighbourhood change.

Compared with the original PACER study, which identified four dimensions (Feelings, Built Environment, Social Environment and Affordability), the Portuguese adaptation yielded a more parsimonious three-factor structure. The original Feelings factor was largely preserved, suggesting that emotional responses to neighbourhood change are stable across cultural contexts. In contrast, the original Built Environment and Social Environment dimensions merged into a single factor reflecting Built, social and economic changes, suggesting that Portuguese respondents perceive physical, commercial and social transformations as interconnected aspects of the same urban process. Additionally, items from the Feelings, Social Environment and Affordability domains clustered into a third factor representing Displacement concerns and neighbourhood disruption, highlighting residential stability and neighbourhood identity as a distinct underlying dimension in the Portuguese context.

Despite the reduction in the number of dimensions, the PACER-PT demonstrated satisfactory psychometric performance. All three factors showed acceptable internal consistency (Cronbach’s alpha=0.70–0.78), supporting the reliability of the adapted instrument. Furthermore, the pattern of correlations with contextual indicators of neighbourhood change and health-related outcomes was conceptually coherent. The factor reflecting “Built, social and economic changes” was most strongly associated with the perceived amount and pace of neighbourhood change, whereas “Positive attitude towards neighbourhood changes” was positively associated with self-rated quality of life and health, providing additional evidence of construct and criterion-related validity.

The comparison of PACER-PT factor scores across participant characteristics provided further support for the instrument’s construct validity. “Positive attitude towards neighbourhood changes” (Factor 2) varied according to age, household income, residential satisfaction and intended length of residence, with more favourable perceptions generally reported by younger participants, those with higher incomes, and respondents expressing greater satisfaction with and stronger intentions to remain in their neighbourhood. “Built, social and economic changes” (Factor 1) differed only by educational level, whereas “Displacement concerns and neighbourhood disruption” (Factor 3) was primarily associated with housing tenure, residential satisfaction and future residential intentions. Overall, contextual and residential characteristics explained greater variation in PACER-PT scores than most sociodemographic characteristics, supporting the instrument’s ability to discriminate between groups expected to differ in their perceptions of neighbourhood change.

The observed differences in PACER-PT factor scores provide evidence of the instrument’s construct validity by showing that perceptions of neighbourhood change vary according to relevant sociodemographic and residential characteristics. Younger participants were more likely to report positive attitudes about neighbourhood changes than older adults. This finding may reflect greater openness to urban transformation and a higher perceived ability to benefit from neighbourhood improvements, as younger adults are often more closely aligned with the lifestyles, amenities, and housing preferences associated with processes of “youthification”, which often occur alongside gentrification and involve urban redevelopment increasingly attracting and catering to younger populations [22]. In contrast, older residents may be more sensitive to the disruption of long-established social ties, place attachment, and local identity that can accompany neighbourhood change. This interpretation is consistent with previous studies showing that gentrification may generate social exclusion, loss of familiar social spaces, and diminished sense of belonging among older adults, even in the absence of physical displacement [23]. Furthermore, participants who were more satisfied with their neighbourhood, as reflected by their willingness to recommend it and their intention to remain living there, consistently reported more positive perceptions of change and fewer concerns related to displacement and neighbourhood disruption. However, these relationships may vary according to the nature of the changes and their consequences for residents, including housing affordability and security of tenure [24].

Several strengths of this study should be acknowledged. First, the adaptation process preserved the conceptual integrity of the original PACER instrument while ensuring linguistic and cultural appropriateness for the Portuguese context. Second, the use of exploratory factor analysis enabled an empirical evaluation of the dimensional structure of the Portuguese version, ensuring that the resulting factors reflected meaningful constructs within the study population. Third, the inclusion of contextual indicators of neighbourhood change allowed the assessment of convergent and criterion-related validity, strengthening the evidence supporting the scale’s validity. Moreover, the inclusion of health indicators enabled an initial exploration of the scale’s relevance for urban health research, which represents its intended field of application. Although the sample was not fully representative of the adult population of the Porto and Lisbon Metropolitan Areas, comparison with Census 2021 data indicated a broadly comparable gender distribution. The overrepresentation of younger and more highly educated participants, which is common in voluntary surveys, may limit the generalisability of the findings. However, the inclusion of participants with diverse sociodemographic and residential characteristics provides support for the applicability of the PACER-PT across a broad range of urban residents. Despite these strengths, several limitations should be considered when interpreting the findings.

Although PACER-PT captures residents’ retrospective perceptions of neighbourhood change, which are relevant, the cross-sectional design does not allow us to assess how these perceptions evolve over time. Longitudinal studies would provide valuable insights into how perceptions evolve over time and how they relate to changes in health outcomes. Additionally, the use of online data collection methods may have introduced selection bias, potentially limiting participation among individuals with lower digital literacy or restricted internet access [25]. Future studies should consider mixed-mode data collection strategies and/or correction strategies to enhance representativeness. Another limitation relates to the geographic scope of the study, which included only residents of the Metropolitan Areas of Porto and Lisbon. While these cities represent major urban centres undergoing rapid transformation, the findings may not be fully generalizable to smaller cities or rural settings [26].

In conclusion, the findings of this study support the reliability and validity of the Portuguese version of PACER as a multidimensional instrument for assessing neighbourhood changes in urban contexts. The PACER-PT demonstrates robust psychometric properties and represents a valuable tool for advancing research on the health impacts of gentrification, touristification, and broader processes of urban transformation in Portugal.

## Supporting information

Supp File 2 (English translation)

## Data Availability

Data produced in the present study are available upon reasonable request to the authors.

