## Supplementary material for "Validation of the PACER-PT: a measure of health-relevant perceptions of gentrification-related neighbourhood change": Supp File 2

### Supplementary File S2

### Perceções sobre Alterações na Área de Residência (PACER-PT)

#### Tipos de Mudança na Área de Residência

Pensando em mudanças ocorridas na sua área de residência, por favor indique em que grau as seguintes mudanças estão a acontecer. Ao responder a estas perguntas, pense sobre o modo como a sua área de residência se transformou durante os últimos três a cinco anos ou como se está a transformar atualmente.

| | Não está a ocorrer | Ocorre pouco | Ocorre bastante | Não sabe |
|---|---|---|---|---|
| 1. Estão a abrir novos estabelecimentos comerciais. | -1 | 1 | 2 | 0 |
| 2. Estabelecimentos comerciais de há longa data estão a ser substituídos por estabelecimentos diferentes. | -1 | 1 | 2 | 0 |
| 3. Estão a abrir mercearias mais caras ou mais sofisticadas. | -1 | 1 | 2 | 0 |
| 4. O custo da habitação (para alugar ou comprar) aumentou. | -1 | 1 | 2 | 0 |
| 5. Houve construção de novos edifícios em lotes vazios ou para substituir edifícios antigos. | -1 | 1 | 2 | 0 |
| 6. Houve construção de novas infraestruturas ou houve melhoria das infraestruturas existentes, tais como parques, ciclovias, transportes ou passeios. | -1 | 1 | 2 | 0 |
| 7. Há quem esteja a comprar imóveis e a renová-los para depois alugar ou vender a preços mais altos. | -1 | 1 | 2 | 0 |
| 8. As transformações estão a gerar tensões ou conflitos entre mim e os meus vizinhos. | 1 | -1 | -2 | 0 |
| 9. Há novas pessoas a mudarem-se para a minha área de residência. | -1 → Q13 | 1 | 2 | 0 |

*Filtro da pergunta 9: se Q9 = -1 (não está a ocorrer), passar para a pergunta 13; se Q9 = 1 ou 2, responder às perguntas 10, 11 e 12.*

**10. Em que aspetos considera que as pessoas que se estão a mudar para a sua área de residência são diferentes de si? Assinale todas as que sejam aplicáveis.**

☐ a. Raça ou origem étnica
☐ b. Nacionalidade ou país de origem
☐ c. Riqueza ou rendimentos
☐ d. Profissão/emprego
☐ e. Escolaridade
☐ f. São estudantes
☐ g. Estrutura familiar
☐ h. Idade
☐ i. Religião
☐ j. Cultura e valores
☐ k. As atividades que gostam de fazer
☐ l. Outro, por favor especifique: ___________________________________
☐ m. Nenhuma; as pessoas que se mudam para a minha área de residência são como eu.

**11. As pessoas que se mudam para a sua área de residência referem-se a esta área usando um nome diferente do nome usado pelos residentes de longa data?**

☐ a. Sim
☐ b. Não
☐ c. Não sei

**12. Até que ponto as pessoas que se mudam para a sua área de residência se interessam pelas instituições e organizações locais existentes (ex.: igrejas, associações locais, escolas)?**

☐ a. Interessam-se muito
☐ b. Interessam-se moderadamente
☐ c. Não se interessam
☐ d. Não aplicável
☐ e. Não sei

#### Mudanças Globais

**13. Numa escala de 1 a 10, sendo que 10 corresponde a grande transformação e 1 a transformação ligeira, no geral, quanto mudou a sua área de residência nos últimos três a cinco anos?**

| 1 | 2 | 3 | 4 | 5 | 6 | 7 | 8 | 9 | 10 |
|---|---|---|---|---|---|---|---|---|---|

*Transformação ligeira* *Grande transformação*

**14. Numa escala de 1 a 10, sendo que 10 corresponde a "rápida" e 1 a "lenta", com que velocidade aconteceram as mudanças na sua área de residência nos últimos três a cinco anos?**

| 1 | 2 | 3 | 4 | 5 | 6 | 7 | 8 | 9 | 10 |
|---|---|---|---|---|---|---|---|---|---|

*Lenta* *Rápida*

**15. Na sua opinião, qual o fator mais importante que está a impulsionar as mudanças na sua área de residência? Selecione um.**

☐ a. Associações locais (ex.: grupos comunitários e entidades sem fins lucrativos)
☐ b. Empresas e estabelecimentos comerciais
☐ c. Universidades, hospitais ou instituições privadas
☐ d. Governo central
☐ e. Autarquias locais (ex.: juntas de freguesia, câmara municipal)
☐ f. Mudanças nas infraestruturas (ex.: requalificação urbana, melhoria de espaços públicos)
☐ g. Transportes novos ou modificados (ex.: novas linhas de transporte público, ciclovias)
☐ h. Construção civil e setor imobiliário
☐ i. Mudanças nas áreas residenciais vizinhas
☐ j. Qualidade das escolas
☐ k. Turismo
☐ l. Outro (por favor indicar): ___________________________

#### Sentimentos sobre as Mudanças

O próximo conjunto de perguntas é sobre o que sente em relação a quaisquer mudanças dentro da sua área de residência. Classifique o quanto concorda ou discorda de cada afirmação, desde "concordo totalmente" a "discordo totalmente".

| | **Concordo totalmente** | **Concordo** | **Não concordo nem discordo** | **Discordo** | **Discordo totalmente** | **Não aplicável** |
|---|---|---|---|---|---|---|
| 16. Se tivesse de mudar de casa agora, eu teria capacidade económica para me mudar para uma casa ou apartamento similar dentro da minha área de residência. | 2 | 1 | 0 | -1 | -2 | 0 |
| 17. Sinto-me bem-vindo/a na maioria dos novos estabelecimentos comerciais da minha área de residência. | 2 | 1 | 0 | -1 | -2 | 0 |
| 18. Sinto que o carácter da minha área de residência mudou. | -2 | -1 | 0 | 1 | 2 | 0 |

| | Concordo totalmente | Concordo | Não concordo nem discordo | Discordo | Discordo totalmente | Não aplicável |
|---|---|---|---|---|---|---|
| 19. Confio nas pessoas que se mudam para a minha área de residência. | 2 | 1 | 0 | -1 | -2 | 0 |
| 20. Sinto-me bem em relação às mudanças a ocorrer na minha área de residência. | 2 | 1 | 0 | -1 | -2 | 0 |
| 21. Tenho receio de ser empurrado/a ou forçado/a a mudar-me para fora da minha área de residência. | -2 | -1 | 0 | 1 | 2 | 0 |
| 22. Apoiaria mudanças na minha área de residência (ex.: novas lojas, passeios, jardins) mesmo que essas mudanças significassem que seria mais caro para mim viver aqui. | 2 | 1 | 0 | -1 | -2 | 0 |
| 23. As mudanças na minha área de residência são destinadas a pessoas como eu. | 2 | 1 | 0 | -1 | -2 | 0 |
| 24. As mudanças que estão a ocorrer na minha área de residência fazem-me duvidar se poderei continuar a viver aqui. | -2 | -1 | 0 | 1 | 2 | 0 |
| 25. Sinto que a minha opinião é ouvida nas decisões sobre as mudanças que ocorrem na minha área de residência. | 2 | 1 | 0 | -1 | -2 | 0 |

#### Área de Residência e Habitação

Agora pedia-lhe que nos fornecesse alguma informação contextual em relação à sua experiência na sua área de residência atual.

**1. Há quanto tempo mora nesta área de residência?**

Anos: _____________

**2. Quais destas razões foram mais importantes para decidir viver nesta área de residência? Escolha as três mais importantes.**

☐ a. Para morar perto de família e amigos
☐ b. Para estar perto do trabalho
☐ c. Acesso a bens e serviços, como lojas, cafés e outros serviços de proximidade
☐ d. Estar perto de transportes públicos
☐ e. Escolas para os meus filhos ou netos
☐ f. Acesso a oportunidades de emprego
☐ g. Segurança da área de residência
☐ h. Custo acessível da habitação
☐ i. Cresci aqui e sempre quis voltar para aqui
☐ j. Vivi sempre aqui
☐ k. Sem escolha/Não tinha outro sítio para ir
☐ l. Outra razão, especifique: ________________________

**3. Por favor, ordene por importância (da mais importante para a menos importante) as razões que o/a levaram a escolher viver nesta área de residência.**

*Opções: as três razões selecionadas na pergunta anterior.*

1.ª ________________________
2.ª ________________________
3.ª ________________________

**4. Neste preciso momento, qual é a probabilidade de recomendar a sua área de residência como um bom sítio para viver?**

☐ a. Muito provável
☐ b. Algo provável
☐ c. Algo improvável
☐ d. Muito improvável

Agora, gostaríamos de lhe fazer algumas perguntas sobre a sua casa atual. Se já viveu nesta área de residência antes, mas numa outra casa, por favor, responda apenas relativamente à sua casa atual.

**5. Há quanto tempo vive na sua casa atual?**

Anos: _____________

**6. Se pudesse escolher, quanto tempo continuaria a morar na sua casa atual?**

☐ a. Mudava-me já
☐ b. Menos de 3 anos
☐ c. Entre 3 e 5 anos
☐ d. Mais de 5 anos mas menos de 10
☐ e. 10 anos ou mais

**7. Atualmente aluga a casa onde mora, é proprietário/a ou outro?**

☐ a. Alugo
☐ b. Proprietário/a
☐ c. Outro, especifique: ________________________
