## Supplementary material for "Validation of the PACER-PT: a measure of health-relevant perceptions of gentrification-related neighbourhood change": Supp File 1

Supplementary File 1, Table 1 – Sociodemographic characteristics of the PACER sample compared with the 2021 Census population of the Porto and Lisbon Metropolitan Areas

| Category | PACER (n) | PACER (%) | Census 2021 (n) | Census (%) |
|---|---|---|---|---|
| **Gender** | **255** | | **3 833 037** | |
| Female | 124 | *48.6* | 2 056 234* | *53.6* |
| Male | 130 | *51.0* | 1 776 803* | *46.4* |
| Other | 1 | *0.4* | *NA* | *NA* |
| **Age Group** | **255** | | **3 833 037** | |
| 18-34 | 85 | *33.3* | 880 457 | *23.0* |
| 35-44 | 66 | *25.9* | 647 892 | *16.9* |
| 45-64 | 90 | *35.3* | 1 303 315 | *34.0* |
| ≥65 | 14 | *5.5* | 1 001 374 | *26.1* |
| **Educational level** | **255** | | **4 606 436**** | |
| Higher education | 135 | *52.9* | 973906** | *21.1* |
| Secondary education | 95 | *37.3* | 1049454** | *22.8* |
| Up to lower secondary | 25 | *9.8* | 2583076** | *56.1* |

*The Portuguese census has no question about gender, so for this purpose, statistics correspond to the variable sex.

**Educational level was derived from the total Census population, as age-specific (≥18 years) educational data were not available.
