## Supplementary material for "Validation of the PACER-PT: a measure of health-relevant perceptions of gentrification-related neighbourhood change": Supp File 2 (English translation)

### Supplementary File S2

### Perceptions of Changes in the Residential Area (PACER-PT)

#### Types of Change in the Residential Area

Thinking about changes that have taken place in your residential area, please indicate the extent to which the following changes are occurring. When answering these questions, think about how your residential area has changed over the past three to five years or how it is changing currently.

| | Not occurring | Occurs a little | Occurs a lot | Don't know |
|---|---|---|---|---|
| 1. New businesses are opening. | -1 | 1 | 2 | 0 |
| 2. Long-standing businesses are being replaced by different businesses. | -1 | 1 | 2 | 0 |
| 3. More expensive or upscale grocery stores are opening. | -1 | 1 | 2 | 0 |
| 4. The cost of housing (renting or buying) has increased. | -1 | 1 | 2 | 0 |
| 5. New buildings have been constructed on vacant lots or to replace old buildings. | -1 | 1 | 2 | 0 |
| 6. New infrastructure has been built or existing infrastructure has been improved, such as parks, cycle lanes, public transport or pavements. | -1 | 1 | 2 | 0 |
| 7. People are buying properties and renovating them in order to rent or sell them at higher prices. | -1 | 1 | 2 | 0 |
| 8. The changes are creating tensions or conflicts between me and my neighbours. | 1 | -1 | -2 | 0 |
| 9. New people are moving into my residential area. | -1 → Q13 | 1 | 2 | 0 |

*Question 9 filter: if Q9 = -1 (not occurring), go to Question 13; if Q9 = 1 or 2, answer Questions 10, 11 and 12.*

**10. In what ways do you think the people moving into your residential area are different from you? Select all that apply.**

☐ a. Race or ethnic origin
☐ b. Nationality or country of origin
☐ c. Wealth or income
☐ d. Occupation/employment
☐ e. Education level
☐ f. They are students
☐ g. Family structure
☐ h. Age
☐ i. Religion
☐ j. Culture and values
☐ k. The activities they enjoy
☐ l. Other, please specify: ____________________________
☐ m. None; the people moving into my residential area are like me.

**11. Do the people moving into your residential area refer to this area using a different name from the one used by long-term residents?**

☐ a. Yes
☐ b. No
☐ c. I don't know

**12. To what extent are the people moving into your residential area interested in existing local institutions and organizations (e.g., churches, local associations, schools)?**

☐ a. Very interested
☐ b. Moderately interested
☐ c. Not interested
☐ d. Not applicable
☐ e. I don't know

#### Overall Changes

**13. On a scale of 1 to 10, where 10 represents major change and 1 represents minor change, overall, how much has your residential area changed over the past three to five years?**

| 1 | 2 | 3 | 4 | 5 | 6 | 7 | 8 | 9 | 10 |
|---|---|---|---|---|---|---|---|---|---|

*Minor change* *Major change*

**14. On a scale of 1 to 10, where 10 represents "fast" and 1 represents "slow", how quickly have changes in your residential area occurred over the past three to five years?**

| 1 | 2 | 3 | 4 | 5 | 6 | 7 | 8 | 9 | 10 |
|---|---|---|---|---|---|---|---|---|---|

*Slow* *Fast*

**15. In your opinion, what is the most important factor driving changes in your residential area? Select one.**

☐ a. Local associations (e.g., community groups and non-profit organizations)
☐ b. Businesses and commercial establishments
☐ c. Universities, hospitals or private institutions
☐ d. Central government
☐ e. Local authorities (e.g., parish councils, municipal councils)
☐ f. Infrastructure changes (e.g., urban regeneration, improvements to public spaces)
☐ g. New or modified transport options (e.g., new public transport routes, cycle lanes)
☐ h. Construction and real estate sector
☐ i. Changes in neighbouring residential areas
☐ j. Quality of schools
☐ k. Tourism
☐ l. Other (please specify): ____________________

#### Feelings About the Changes

The next set of questions concerns how you feel about any changes in your residential area. Please rate how much you agree or disagree with each statement, from "strongly agree" to "strongly disagree".

| | Strongly agree | Agree | Neither agree nor disagree | Disagree | Strongly disagree | Not applicable |
|---|---|---|---|---|---|---|
| 16. If I had to move house now, I would be financially able to move to a similar house or apartment within my residential area. | 2 | 1 | 0 | -1 | -2 | 0 |
| 17. I feel welcome in most of the new businesses in my residential area. | 2 | 1 | 0 | -1 | -2 | 0 |
| 18. I feel that the character of my residential area has changed. | -2 | -1 | 0 | 1 | 2 | 0 |
| 19. I trust the people moving into my residential area. | 2 | 1 | 0 | -1 | -2 | 0 |
| 20. I feel positive about the changes taking place in my residential area. | 2 | 1 | 0 | -1 | -2 | 0 |

| | Strongly agree | Agree | Neither agree nor disagree | Disagree | Strongly disagree | Not applicable |
|---|---|---|---|---|---|---|
| 21. I am afraid of being pushed or forced to move out of my residential area. | -2 | -1 | 0 | 1 | 2 | 0 |
| 22. I would support changes in my residential area (e.g., new shops, pavements, gardens), even if those changes meant it would be more expensive for me to live here. | 2 | 1 | 0 | -1 | -2 | 0 |
| 23. The changes in my residential area are intended for people like me. | 2 | 1 | 0 | -1 | -2 | 0 |
| 24. The changes taking place in my residential area make me doubt whether I will be able to continue living here. | -2 | -1 | 0 | 1 | 2 | 0 |
| 25. I feel that my opinion is heard in decisions about the changes taking place in my residential area. | 2 | 1 | 0 | -1 | -2 | 0 |

#### Residential Area and Housing

We would now like you to provide some contextual information about your experience in your current residential area.

**1. How long have you lived in this residential area?**

Years: ______________

**2. Which of the following reasons were most important in your decision to live in this residential area? Choose the three most important.**

☐ a. To live close to family and friends
☐ b. To be close to work
☐ c. Access to goods and services, such as shops, cafés and other nearby services
☐ d. To be close to public transport
☐ e. Schools for my children or grandchildren
☐ f. Access to employment opportunities
☐ g. Safety of the residential area
☐ h. Affordable housing costs
☐ i. I grew up here and have always wanted to return
☐ j. I have always lived here
☐ k. No choice/I had nowhere else to go
☐ l. Other reason, please specify: ______________________

**3. Please rank in order of importance (from most important to least important) the reasons that led you to choose to live in this residential area.**

*Options: the three reasons selected in the previous question.*

1st __________________________
2nd __________________________
3rd __________________________

**4. At this moment, how likely are you to recommend your residential area as a good place to live?**

☐ a. Very likely
☐ b. Somewhat likely
☐ c. Somewhat unlikely
☐ d. Very unlikely

We would now like to ask you some questions about your current home. If you have lived in this residential area before, but in a different home, please answer only with regard to your current home.

**5. How long have you lived in your current home?**

Years: ______________

**6. If you could choose, how much longer would you continue living in your current home?**

☐ a. I would move out now
☐ b. Less than 3 years
☐ c. Between 3 and 5 years
☐ d. More than 5 years but less than 10
☐ e. 10 years or more

**7. Do you currently rent the home you live in, own it, or have another arrangement?**

☐ a. I rent
☐ b. I own it
☐ c. Other, please specify: ______________________
